# Prevalence and Correlates of Diabetes: Inception Cohort of a Semi-Urban Population of Southern India

**DOI:** 10.1101/2020.08.21.20179739

**Authors:** Mandeep Singh, Atul Kotwal, Chetan Mittal, PC Reddy, J Subbanna, Ram Babu, Anupam Kotwal

## Abstract

**Introduction:** India has the second highest number of people with diabetes, however few studies assessed true burden and ensured screening of pre-diabetics. This cross sectional survey (inception cohort) was conducted to estimate the burden and correlates of one of the common risk factors (hyperglycemia / diabetes) likely to impact the morbidity and mortality due to Non-communicable diseases.

**Research Design and Methods:** A cross sectional survey was conducted in villages under Chittoor District, Andhra Pradesh, India. 15,888 individuals (62.1% females and 37.9% males) above 15 years of age participated in the study.

**Results:** 960 (6.04%) had a history of diabetes while 592 (3.67%) had high random capillary blood glucose (RCBG) ≥200 mg/dL. HbA1c and fasting blood sugar was tested in a subset of those with high RCBG (n = 248) but with no self-reported diabetes and 212 (85.48%) had HbA1c levels > 6.5 and fasting blood sugar ≥126 mg/dL. Thus, 85.48% (n = 506) with high RCBG were likely to be diabetic. The likely magnitude of diabetes in the community was 9.23%. Another 2,331 (14.67%) were found to be at a higher risk of developing diabetes (RCBG 140 to 200 mg/dL). Receiver Operator Characteristic analysis revealed waist circumference as the most important anthropometric measurement predicting high RCBG. Comparison of diabetes and non-diabetics by multivariate logistic regression showed that male gender, age, weight, history of hypertension, family history of diabetes and waist circumference played a significant role in the prevalence of diabetes.

**Conclusions:** There is high prevalence of diabetes in the rural and semi-urban India. Waist Circumference was the best predictor of Diabetes. RCBG can be considered as a easy, cheap, reliable and important tool for community diagnosis and management of diabetes.

**Strengths and limitations of the study:**

- A large planned cross-sectional survey to establish a cohort with community participation
- Quality assurance and control measures at all levels for high internl validity
- Follow up assurance for the entire community
- These results are from Cross Sectional survey data and thus provides only associations
- Generealizeability to similar populations

*What is already known?:*

- Global burden of disease projected India to have a manifold increase in diabetes between 1995–2025.
- Diabetes threat is being under-prioritised with increasing pool of undiagnosed diabetes.

*What are the new findings?:*

- Total prevalence of Diabetes 9.77% and 14.67% were at risk of developing Diabetes
- 85.5 % individuals with RCBG≥200mg/dL conform to diabetes as per ADA criteria.
- Waist Circumference was the best predictor of Diabetes.

*What do the new findings imply?:*

- RCBG can be a reliable, cheap and important tool for community diagnosis and management of diabetes.
- Cohort established, interventions being implememted with follow up.

## Introduction

India is in an epidemiological transition phase between communicable and non-communicable diseases (NCDs). Among the NCDs, diabetes is in a potentially epidemic state and this has attracted a lot of attention in recent years. It has been reported that India has the second highest number of people with diabetes (69.2 million) and will have 123.5 million persons with diabetes by 2040, as per projections.[1] A recent study showed the overall prevalence of diabetes as 7.3% across 15 states of India; with higher prevalence in Andhra Pradesh (8.4% overall;12.6% in urban and 6.3% in rural areas).[2] Another one reported prevalence of 13.7 % in urban and 7.8% in rural areas of Tamil Nadu.[3] The National Urban Survey (2001) reported age and gender standardized prevalence of 13.5%, 12.4% and 16.6% in Chennai, Bangalore and Hyderabad, respectively.[4] Earlier, the global burden of disease study projected India to have manifold increase in diabetes between 1995-2025, with hisher prevalence among middle-aged individuals similar to the developed countries.[5]

Due to simplicity of implementation and cost, community surveys for diabetes often use random capillary blood glucose (RCBG). There is no consensus statement for RCBG cut-off’s in Indian population, however as per American Diabetes Association (ADA) guidelines, RCBG ≥ 200mg/dL is considered diagnostic if accompanied by symptoms of hyperglycemia.[6] RCBG range of 140-199 mg/dL has often been referred to as prediabetes in many studies, though the ideal level for carrying out further screening with HbA1c/OGTTs, is still in debate, with some studies recommending screening at RCBG > 108-110 mg/dLand others recommending > 120 mg/dL, to avoid additional costs with few advantages.[7]

Among the correlates for diabetes, high body mass index (BMI) has been accorded relatively less importance for South Asians including Indians especially because of its inability to differentiate between fat and fat-free mass.[8] Additionally, BMI underestimates visceral fat particulary in South Asians.[9] A consensus statement was released for Indians with different cut-offs for BMI and waist circumference (WC).[10] Overweight was defined as BMI 23-24.9 kg/m^2^ and obesity as ≥25 kg/m^2^ as compared to higher values of internationally accepted cut-offs of 25–29.9 kg/m^2^ and ≥30 kg/m^2^ for overweight and obesity, respectively. Similarly, WC cut-offs were defined as – Action level 1: men-78 cm, women-72 cm; and Action level 2: men-90 cm, women-80 cm. Other than anthropometric measurements, few studies have also correlated diabetes with occupation, income and education. A study to analyze the cumulative incidence and relative risk of type 2 diabetes found significantly lower prevalence among farmers.[11] Another recent study from North India did not find any significant impact of education but found significantly more odds of developing diabetes among the married, divorced, separated and widowed individuals as compared to unmarried individuals in a multivariate regression model analysis.[12]

While efforts by various governments and agencies in the developed world have resulted in positive health outcomes for their communities, studies assessing the true burden of diabetes and efforts for ensuring screening of the pre-diabetes and enrolling in a preventive program in India have been sparse. The diabetes threat is thereby, being under-prioritised and the pool of undiagnosed diabetes, continues to grow along with the complications due to the uncontrolled hyperglycaemia.

This study was conducted in the Indian state of Andhra Pradesh in South India, in a semi urban population to: estimate baseline diabetes prevalence as part of inception cohort; identify undiagnosed diabetes (RCBG≥ 200 mg/dL and unaware of diabetes status); identify those at a higher risk of developing diabetes and probable prediabetes (RCBG between 140-199 mg/dL); identify the correlates among these groups; enroll persons with diabetes (self reported) under a regulated monitoring in the first phase by instituting management and monitoring their progress regularly and also behaviour change communication (BCC). The cohort is being followed up and as a part of protocol, the newly diagnosed persons with diabetes and those at a higher risk of developing diabetes are being assessed by carrying out HBA1c and oral glucose tolerance tests in accordance with WHO criteria and managed.

## Methods and Materials

A cross-sectional survey was conducted using modified WHO STEPS questionnaire in a group of villages under the Thavanampalle *Mandal* - one of the 66 *Mandals* in Chittoor District in the South Indian state of Andhra Pradesh covered under ‘Total Health’, a CSR arm of Apollo Hospitals Enterprises Ltd and with a view to establish a cohort. As per 2011 census, Thavanampalle Mandal comprises 32 Gram Panchayats and 195 villages with a total population of 53,708 (49.7% males and 50.3% females).

The study was approved by the IEC of Apollo Health Education and Research Foundation (AHERF), Apollo Hospitals, Jubilee Hills, Hyderabad. Informed consent was obtained from all participants in a language they understood (Telugu/Kannada) and signature/thumb imprint obtained. In phase I, 98 villages were selected by stratified random sampling, with a total population of 27,483 and family size of 4.04 as per the census of 2011. Of these, repeated household visits ensured enrolment of 22,303 (81.3%). Data were collated and analysed for 15,888 individuals (62.1% females and 37.9% males) above 15 years of age from 8947 families with an average number of 1.86 members from each family (median 2). Trained healthcare workers keyed in the data in android tablets using application software specifically developed for this survey. Quality assurance measures included training of data collectors and supervision of a proportion of their visits. RCBG was measured with a glucometer (Accu Chek Performa). Calibration of glucometers was carried out every month by randomly matching 2% of the blood glucose level result with the laboratory at the local Apollo hospital and every week by comparing with another glucometer. Results within 15% of the laboratory reading were considered accurate.

HbA1c and fasting blood sugar levels were tested in 248 individuals with RCBG ≥200 mg/dL. Urine examination for sugar and protein was done at the time of the survey. Individuals with history of diabetes and those with RCBG≥140 mg/dL are being further followed up by investigations, including HbA1c, serum creatinine, blood urea and eye examination at the Foundation’s clinics established in the project area. Cut-offs for BMI and waist circumference were taken as per consensus statement for analysis.[10]

A database was created in My SQL and analysed using iStata and SPSS 16. Kolmogorov–Smirnov test was used as test of normality. Statistical analysis of independent categorical variables with dependent continuous data was done using t-test, analysis of variance (ANOVA) and correlation coefficient. Univariate and multivariate regression models were utilized for risk factor analysis. Further details of model have been provided in results section.

### Patient and Public Involvement statement

The entire population of the selected area was informed about the inception cohort and baseline survey. The potential participants were informed regrading the components of the baseline survey, which were: collection of information about socio-demographic variables; anthropometry; bmedical checkup including general physical examination; blood tests for biochemistry and hematology only. They were also informed that blood samples will not be used for any genetic studies or biobanking. The community was also informed regarding treatment offered at nominal charges for ailments diagnosed during the survey and also the health education to be imparted. The informed consent was then obtained from the community leaders and also from each participant. The results were shared with respective participants confidentially and all participants are being followed up regularly. Thus this project is being conducted in the true spirit of community participation, social mobilization within the principles of ehtics for biomedical research involving human participants.[13]

## Results

The prevalence of known diabetes (self-reported during questionnaire response) (Table 1) was 6.04% (N = 960). Another 592 (3.73%) had RCBG ≥200mg/dL. Additionally, 2,331 (14.67%) were at high risk of developing diabetes (RCBG = 140–199 mg/dL). The overall prevalence of diabetes was 9.77% (95% CI, 9.31, 10.24).

**Table 1:** Prevalence of Diabetes in the surveyed population.

| Sr No | Category/Group | N (%) | 95 % CI | Classification |
| --- | --- | --- | --- | --- |
| 1 | Normal | 12,005 (75.56) | 74.88, 76.22 | Non – Diabetic |
| 2 | Newly diagnosed “High Risk of developing Diabetes” (RCBG 140-199 mg/dL) | 2,331 (14.67) | 14.12, 15.23 |  |
| 3 | Newly Identified Diabetics (RCBG≥200mg/dL) | 592 (3.73) | 3.44, 4.03 | Diabetic |
| 4 | Known Diabetics (Self reported) | 960 (6.04) | 5.68, 6.42 |  |
| 5 | Total | 15,888 (100.0) |  |  |

Of the 248 participants with RCBG≥200mg/dL, 85.48 % (95% CI, 80.47, 89.62) had HbA1c levels conforming to diabetes as per ADA criteria and another 12.52% had HbA1c levels between 5.7-6.4 (prediabetes). For purposes of analysis the individuals with self reporting diabetes and those with RCBG≥200 mg/dL were classified as the ‘Diabetes group’ and the others i.e. those not self reporting diabetes during questionnaire response with normal RCBG (< 120 mg/dL) and those with RCBG between 140-199mg/dL were classified as the ‘Non-diabetes group’.

Among those aware of their diabetes, 534 (50.8%) had RCBG≥200 mg/dL, showing poor control, despite knowing their diabetes status and being on management of some kind. Only 26% of these had RCBG < 140 mg/dL.

The mean RCBG level in the surveyed population was 130.1 mg/dL (95%CI: 129.2, 130.9 mg/dL) with males 135.4 (95% CI, 133.8, 136.9 mg/dL) and females 126.8 mg/dL (95% CI, 125.9, 127.8 mg/dL) as shown in Table 2. The variables showing statistically significant association with RCBG were included in the multivariate model. Male sex, rising age, hypertension, increasing BMI, family history of diabetes, increasing waist circumference and weight gain showed significant relationship with increase in RCBG. Family history of diabetes and age were the most significant predictors of RCBG levels (Table 2). Correlation of RCBG with anthropometric variables Showed that correlation was highest with WC (p = 0.001), followed by weight, WHR and BMI. Details provided in Table 2.

**Table 2:**
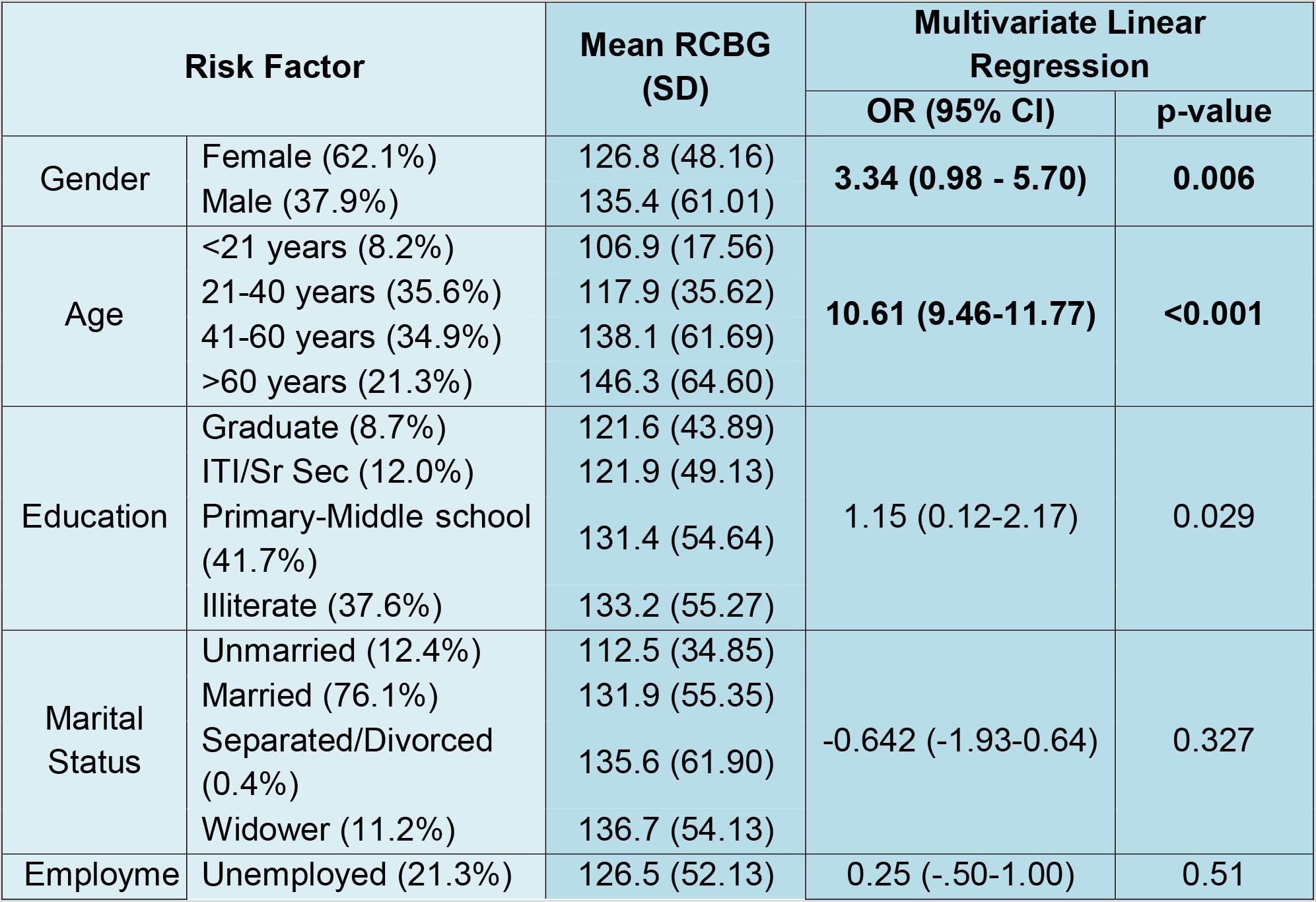
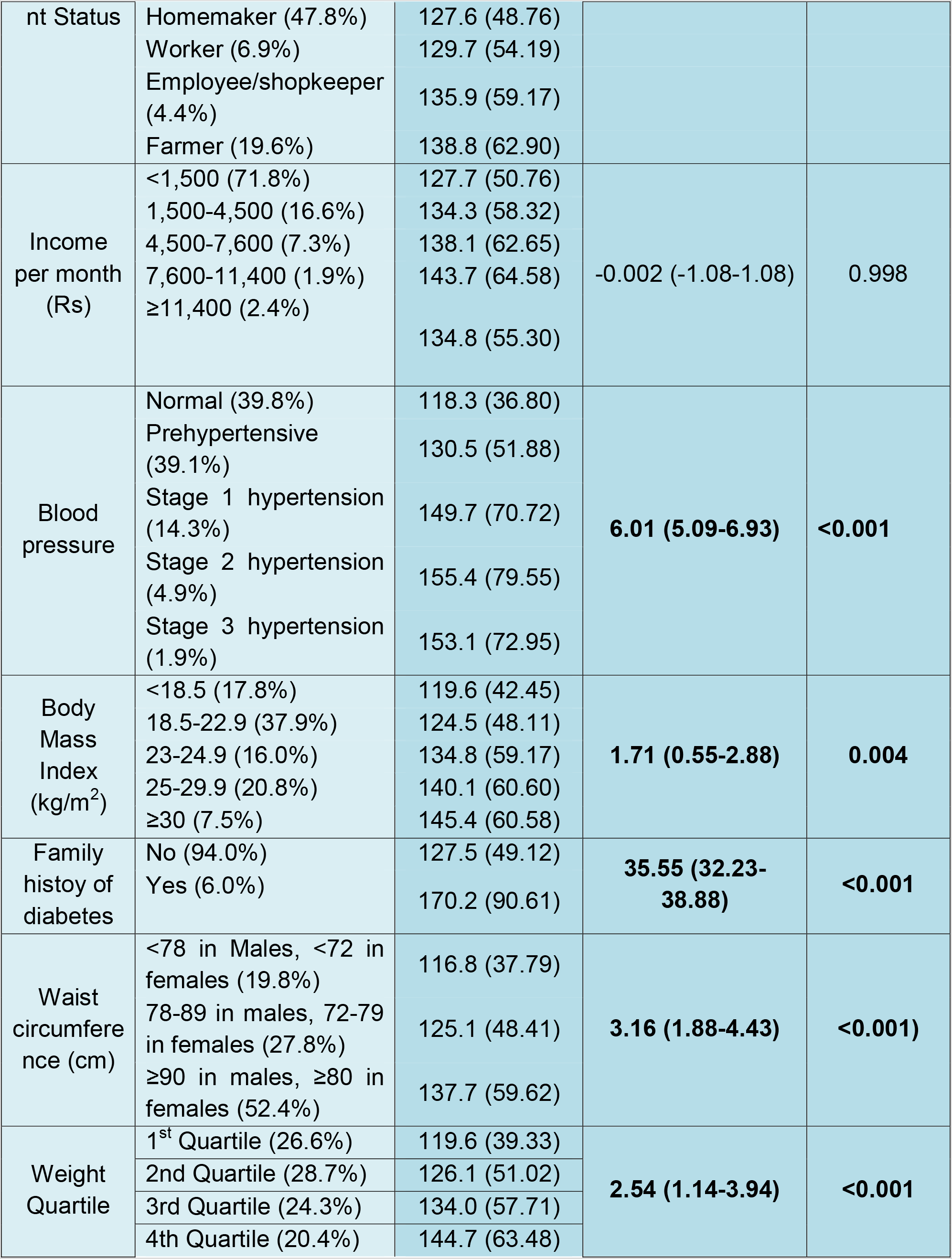
Multivariate linear regression for analyzing various risk factors for RCBG.

The study population was categorised into the diabetes and non-diabetes groups (Table 1). Receiver Operator Characteristic (ROC) analysis (Table 3 and Figure) of anthropometric measurements including WC, hip circumference (HC), BMI, weight, WHR and height showed that WC had the best correlation with Diabetes with area under curve being 0.701 in females and 0.693 in males. Youden’s index showed a threshold of 86.5 cm in females and 89.5 cm in males.

**Table 3:** ROC analysis for various anthropometric measurements.

| Anthropometric parameter | Female |  |  | Male |  |  |
| --- | --- | --- | --- | --- | --- | --- |
|  | Area under curve | 95% CI | p-value | Area under curve | 95% CI | p-value |
| **Waist Circumference** | 0.702 | 0.684-0.720 | <0.001 | 0.693 | 0.673-0.712 | <0.001 |
| **Hip Circumference** | 0.689 | 0.670-0.708 | <0.001 | 0.646 | 0.626-0.667 | <0.001 |
| **Body Mass Index** | 0.679 | 0.661-0.698 | <0.001 | 0.634 | 0.613-0.654 | <0.001 |
| **Weight** | 0.676 | 0.657-0.694 | <0.001 | 0.628 | 0.608-0.649 | <0.001 |
| **Waist Hip Ratio** | 0.564 | 0.544-0.583 | <0.001 | 0.639 | 0.619-0.658 | <0.001 |
| **Height** | 0.522 | 0.501-0.542 | 0.046 | 0.513 | 0.492-0.535 | 0.229 |

Youden's Index for Waist Circumference : Females = 86.5cm (Sensitivity – 67.7, Specificity – 63.2); Males = 89.5cm (Sensitivity – 64.5, Specificity – 64.3)

**Figure:**
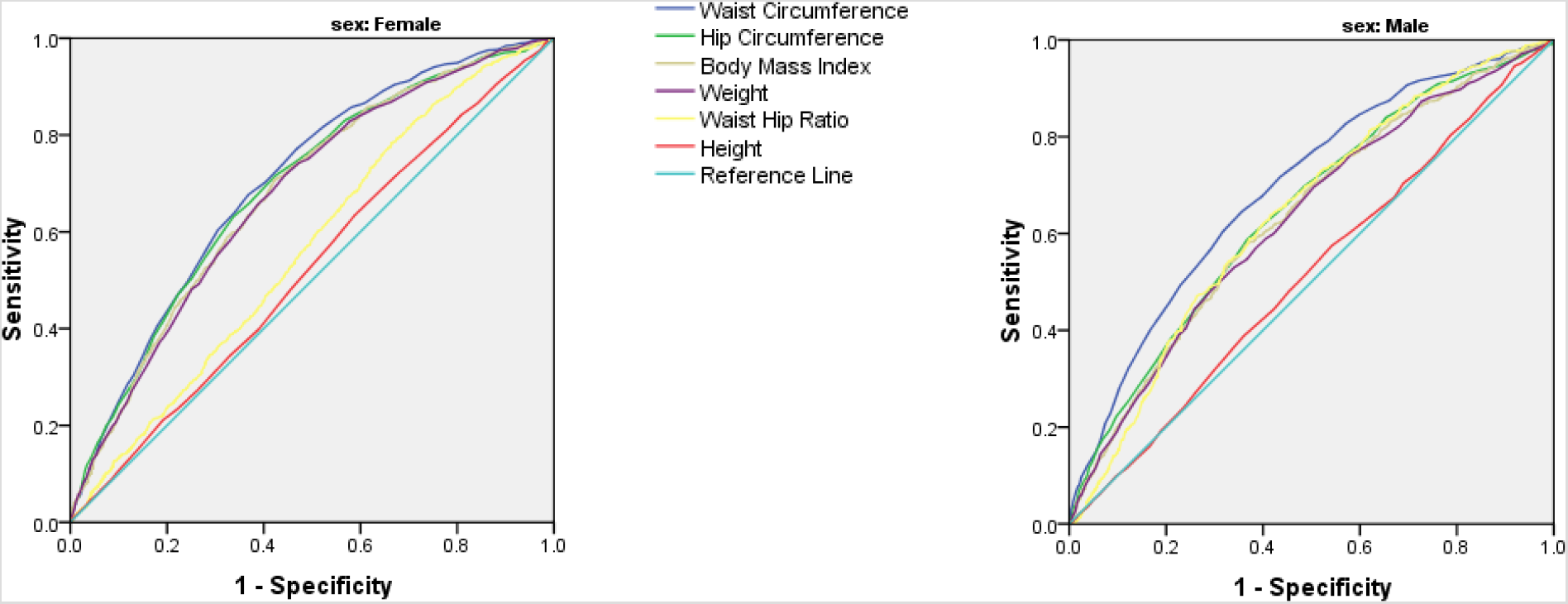
ROC curve analysis for various anthropometric measurements

A univariate followed by multivariate logistic regression was done for risk factors for diabetes and details have been provided in Table 3. Odds of abnormally high RCBG (≥200 mg/dL) increased by 77% in males as compared to females (OR 1.77; 95% CI,1.59, 1.96) and this difference was statistically significant (P < 0.001). Age also showed statistically significant impact on odds of abnormally high RCBG which increased by 4% for every one year increase in age (OR 1.04; 1.04, 1.05) (P < 0.001).

**Table 4:** Multivariate logistic regression for identifying risk factors for Diabetes in study population.

| Risk Factor (%) |  | Prevalence<br>RCBG (%) | Multivariate Logistic Regression |  |
| --- | --- | --- | --- | --- |
|  |  | >200mg/dl | Odds ratio (95% CI) | p-value |
| Gender | Female (62.1) | 7.8 | 1 |  |
|  | Male (37.9) | 13.0 | **1.48 (1.17-1.87)** | **0.001** |
| Age | <21 years (8.2) | 0.5 | 1 |  |
|  | 21-40 years (35.6) | 3.1 | **4.14 (1.76-9.70)** | **0.001** |
|  | 41-60 years (34.9) | 12.9 | **15.39 (6.54-36.23)** | **<0.001** |
|  | >60 years (21.3) | 19.3 | **27.36(11.58-64.65)** | **<0.001** |
| Education | Graduate (8.7) | 7.9 | 1 |  |
|  | ITI/Sr Sec (12.0) | 7.1 | 1.06 (0.78-1.44) | 0.730 |
|  | Primary-Middle school (41.7) | 10.8 | 1.10 (0.85-1.42) | 0.463 |
|  | Illiterate (37.6) | 10.0 | 0.93 (0.71-1.23) | 0.617 |
| Marital Status | Married (76.1) | 10.5 | 1 |  |
|  | Separated/Divorced (0.4) | 6.1 | 0.86 (0.30-2.49) | 0.777 |
|  | Unmarried (12.4) | 2.8 | 0.83 (0.60-1.14) | 0.250 |
|  | Widower (11.2) | 12.6 | 0.95 (0.78-1.15) | 0.596 |
| Employment Status | Employee/shopkeeper (4.4) | 14.4 | 1 |  |
|  | Farmer (19.6) | 13.6 | 0.80 (0.61-1.06) | 0.124 |
|  | Homemaker (47.8) | 8.1 | 0.92 (0.65-1.29) | 0.615 |
|  | Unemployed (21.3) | 9.0 | 0.97 (0.70-1.35) | 0.868 |
|  | Worker (6.9) | 9.9 | 0.77 (0.55-1.07) | 0.118 |
| Income per month (Rs) | <1,500 (71.8) | 8.6 | 1 |  |
|  | 1,500-4,500 (16.6) | 10.9 | 1.00 (0.82-1.22) | 0.998 |
|  | 4,500-7,600 (7.3) | 14.8 | 1.11 (0.86-1.42) | 0.422 |
|  | 7,600-11,400 (1.9) | 20.2 | 1.40 (0.98-2.01) | 0.067 |
|  | ≥11,400 (2.4) | 12.3 | 0.74 (0.51-1.07) | 0.106 |
| Blood pressure | Normal (39.8) | 3.7 | 1 |  |
|  | Prehypertensive (39.1) | 9.5 | **1.46 (1.23-1.72)** | **<0.001** |
|  | Stage 1 hypertension (14.3) | 20.5 | **2.43 (2.02-2.92)** | **<0.001** |
|  | Stage 2 hypertension (4.9) | 23.6 | **2.31 (1.83-2.93)** | **<0.001** |
|  | Stage 3 hypertension (1.9) | 25.9 | **2.89 (2.10-3.97)** | **<0.001** |
| Body Mass Index (kg/m<sup>2</sup>) | <18.5 (17.8) | 3.9 | 1 |  |
|  | 18.5-22.9 (37.9) | 7.1 | 1.03 (0.79-1.34) | 0.836 |
|  | 23-24.9 (16.0) | 11.5 | 1.09 (0.79-1.49) | 0.606 |
|  | 25-29.9 (20.8) | 14.9 | 1.23 (0.89-1.71) | 0.218 |
|  | ≥30 (7.5) | 19.5 | **1.58 (1.08-2.30)** | **0.018** |
| Family history of diabetes | No (94.0) | 7.8 | 1 |  |
|  | Yes (6.0) | 40.8 | **6.99 (5.94-8.22)** | **<0.001** |
| Waist circumference (cm) | <78 in Males, <72 in females (19.8) | 3.1 | 1 |  |
|  | 78-89 in males, 72-79 in females (27.8) | 6.5 | 1.19 (0.92-1.53) | 0.191 |
|  | ≥90 in males, ≥80 in females (52.4) | 14.0 | **1.95 (1.51-2.52)** | **<0.001** |
| Weight Quartile | 1<sup>st</sup> Quartile (26.6) | 3.9 | 1 |  |
|  | 2<sup>nd</sup> Quartile (28.7) | 7.4 | **1.42 (1.12-1.81)** | **0.004** |
|  | 3<sup>rd</sup> Quartile (24.3) | 12.0 | **1.65 (1.25-2.18)** | **<0.001** |
|  | 4<sup>th</sup> Quartile (20.4) | 18.0 | **1.73 (1.25-2.40)** | **0.001** |

Among various occupational categories, employees or shopkeepers had statistically significant higher odds of abnormally high RCBG as compared to worker (OR 1.53; 1.15, 2.05; P = 0.004). Odds of abnormally high RCBG increased statistically significantly with income Rs 7,600-11,400 (OR 2.68; 2.01, 3.57; P < 0.001) and income Rs 4,500-7,600 (OR 1.84; 1.55, 2.20; P < 0.001) when compared to participants with lower income. Odds of abnormally high RCBG increased among widows/widowers as compared to unmarried individuals (OR 1.23; 1.06, 1.43; P = 0.007). Hypertension also emerged as a significant predictor for diabetes (OR 3.93; 95% CI:3.53-4.38; P-value < 0.001). Odds of high RCBG increased with increasing BMI (OR 1.11; 1.10, 1.12; P < 0.001). Higher quartiles of body weight (2nd-4th) showed statistically significant higher risk of diabetes as compared to those in first quartile. Male gender, increasing age, rising blood pressure, waist circumference more than Action 2 cut-offs and increasing weight retained their significance in influencing abnormally high RCBG as outcome.

## Discussion

The overall prevalence of Diabetes in our study (9.77%) was between the urban (12.6%) and rural (6.3%) of Andhra Pradesh,[2] and also of urban (13.7 %) and rural (7.8%) in Tamil Nadu.[3] Our results were almost similar to the prevalence found in North India in a recent study.[12] Our study finding of 50.8 % of diabetics having poor control is lesser than the North Indian study (65%) and a health camp-based study in Chennai, wherein 65.4% of the known case of diabetics had uncontrolled sugar values of > 200 mg/dl.[12.14] Thus, poor glycemic control is a major problem in India. Coupled with this is the fact that even the self-care activities among diabetics have been found to be low in India.[14]

BMI and WHR measures were found as predictors of hypertension in the same population.[16] For diabetes, increasing age, family hisotry, current hypertension, obesity, WC, BMI were found as correlates, Studies in other parts of India have also found similar correlates of Diabetes.[12,17] with only one difference as our study found Waist Circumference (WC) as the best predictor of Diabetes, which is a novel finding for India, although it has been reported in otherspopulations like Mexican Americans, Black Americans, White Americans, Chinese and Africans.[18,19.20,21]

A not significant (P>0.05) association was found among those with BMI > 30kg/m^2^ only. This outcome supports the findings in other studies that BMI may not be the best anthropometric screening mechanism for diabetes, primarily because of its inability to differentiate between fat and fat-free mass. Dominance of waist circumference and weight in risk assessment for diabetes clearly points to importance of visceral fat. WC and BMI have been found to be indicators of prediabetes even in adoloscents.[22] Therefore, robust control strategies need to be thought for the Indian population to bring appropriate lifestyle changes at a young age for checking this rise in visceral fat culminating later into diabetes.

## Conclusion

Concerted efforts are required to prevent, control and manage diabets in our country. RCBG can be considered as a very reliable and important tool for community diagnosis and management strategies for diabetes, specially considering its cost and ease of carrying out in a survey.

## Recommendations

The findings need to be verified in other settings / states of our vast country. The research desingn permissted analysis of correlates. The established cohort is being followed up and future research into causative factors will be carried out. Community wide planned interventions and regular follow up for assessing trends of various diseases, chronic conditions, behaviours and practices are also ongoing.

## Data Availability

Data available on request

## Contributorship

All authors contributed significantly in design of the project (AK, MS); implementation (MS, AK, JS, RB, PSR); field work for data collection (JS, RB) and supervision (MS, PSR); data collation and analysis (CM, AK); manuscript preparation (AK, CM, AK2) and approval (MS, AK).

